# Using the ECHILD Database to Explore Educational and Health Outcomes of Unaccompanied Asylum-Seeking Children living in England (2005 – 2021)

**DOI:** 10.64898/2026.03.04.26347576

**Authors:** Rebecca Langella, Pia Hardelid, Kate Marie Lewis

## Abstract

UK-based quantitative research on the health and education outcomes of Unaccompanied Asylum-Seeking Children (UASC) remains limited, especially at national level. Linked administrative data provide an unprecedented opportunity to study these outcomes among UASC. This paper lays a foundation for further research, particularly examining the influence of socio-demographic, legal and environmental factors on UASC’s health and educational outcomes.

We described the UASC population with a first recorded episode of local authority care between 1st April 2005 and 31st March 2021 in ECHILD, which gathers national records for England, by age, gender, ethnicity, region, and placement type. We calculated linkage rates between the social care and educational dataset, estimating how many UASC were recorded as being enrolled in state-funded schools. We also assessed how many of those linked to the school dataset was linked to National Health Service (NHS) datasets. Finally, we explored how linkage rates between social care, education, and NHS datasets vary by socio-demographic factors and placement type.

There were 37,170 UASC recorded in the ECHILD of which 32,570 (88%) were male and 24,290 (65%) aged 16 – 17 years. We found 7,740 (21%) UASC recorded as being enrolled in state funded schools, of whom 6,690 (88%) were also linked to NHS data. The linkage rate for UASC in the social care to health datasets was therefore 19%. Of those 16–17 years at entry in social care, 4% (1,060/24,290) were recorded as enrolled in school compared to 50% (6,390/12,880) under 16 years.

Linkage to the school, and subsequently to the NHS dataset, wholly depends on enrolled state-funded education, excluding College and Sixth-form education. Despite this limitation, we characterised a national cohort of 6,890 UASC in England whose social care, education, and health outcomes can be examined.

## 1. Introduction & rationale

In the UK, unaccompanied asylum-seeking children (UASC) are individuals under the age of 18 years arriving in the country seeking protection without the care of a parent or legal guardian [1]. Under the Children Act 1989, UASC are recognised as looked-after children, meaning that local authorities act as their corporate parents and are responsible for ensuring they receive the same entitlements and support as any other child in care [2]. This legal duty encompasses the provision of accommodation, care, and support, including free access to education, primary and secondary healthcare services, and legal representation. The number of UASC in the UK has been increasing steadily in recent years, with 7,380 UASC residing under local authority care in 2024, compared with 5,150 in 2019 [3, 4].

Compared with other children in care, UASC face distinct health, social, and educational challenges, shaped both by their country of origin and by the circumstances of their migration journey [5]. Many arrive from low or middle-income countries and may have experienced conflict, persecution, or violence, and long, hazardous journeys to reach the UK [6]. During migration, UASC are often exposed to multiple risks, including deprivation, physical and sexual assault, lack of shelter, and unsafe travel conditions [6]. Post-arrival experiences and ongoing precariousness further shape UASC’s health and overall wellbeing. The detrimental impact of the asylum process on mental health is well documented. A recent systematic and thematic review focusing on the UK and EU+ countries found that prolonged waits for asylum decisions significantly contribute to psychological distress [7, 8].

Despite these challenges, UASC frequently demonstrate considerable resilience, particularly through their engagement in education. International research has identified school attendance as a key protective factor against poor mental health outcomes for UASC, while also supporting social integration, creating stability, and fostering a sense of community [9].

However, quantitative research in the UK on UASC’s social care, health, and educational outcomes remains limited, particularly at the national level, and few studies examine the intersections between these areas. Health research often relies on small, local samples, typically drawn from Initial Health Assessments (IHAs), which are comprehensive screenings conducted before children enter care. Longitudinal or comprehensive physical health data are largely absent [10]. Some studies [11, 10, 12] have used linked social care and educational administrative data to examine UASC educational outcomes, but no studies have yet analysed health and education data together.

Administrative data provide a powerful resource for describing the experiences and health and education outcomes of UASC at a national level. This study uses the Education and Child Health Insights from Linked Data (ECHILD) dataset, which combines routinely collected records from NHS hospital care, local authority social services, and state-funded schools. By linking these datasets, ECHILD could offer a comprehensive, longitudinal view of UASC’s interactions with state education, health, and care systems across England. Such linkage may enable assessment of how placement settings and policy decisions shape UASC’s health and educational trajectories.

However, to realise this potential, it is crucial to understand both the extent of linkage and the factors associated with linkage. This study therefore aims to: (1) describe the socio-demographic characteristics and placement types of UASC recorded in the ECHILD social care dataset, (2) assess linkage rates across social care, educational and healthcare datasets, and (3) examine how these vary by socio-demographic characteristics and placement type.

## 2. Methods

### 2.1. Data sources

#### 2.1.1. ECHILD

CHILD brings together in National Pupil Database (NPD) education datasets, including the Children Looked After (CLA) social care dataset, and NHS health data for young people aged 0-24 years in England who were born between September 1^st^ 1995 and 31^st^ of August 2020 [13]. We used the National Pupil Database (NPD) and Children Looked After (CLA) dataset, respectively, and the ECHILD linkage spine to identify pupils who had any National Health Service (NHS) contact.

The NPD contains individual-level information on pupils in state-funded schools in England, including demographic, educational attainment, special educational needs and disability provision, attendance, exclusions, and the use of children’s social care services [14]. One of the social care modules included in the NPD is the CLA dataset, a national, child-level dataset that records information on all looked after children and recent care leavers in England. The CLA includes data on child characteristics, out-of-home placements, and adoptions.

The basic unit in the CLA is care episodes. A new care episode might be started for several reasons, including, but not limited to, change of placement, of carers or legal status. Data collection for the CLA began in 1992 and has continued through an annual online census conducted by local authorities. This Census is collected on the 31^st^ of March of each year. Since 2004 CLA also includes a flag for whether a child looked after is a UASC.

#### 2.1.2. Linkage across social care, educational and NHS datasets

In ECHILD, children in the CLA dataset must be enrolled in state-funded school in England to be eligible for linkage to the NPD. A child is assigned a Unique Pupil Number (UPN) when they are first enrolled in a state-funded education (nursery or school), which is generated by the school, or in some circumstances by local authorities for children not attached to state school. Since April 2005, local authorities have submitted social care returns to the Department of Education manually including UPNs, where found, enabling linkage to NPD records. Researchers receive only a pseudo-anonymised version of the UPN called the Pupil Matching Reference Number (PMR). Some post-compulsory institutions, such as Further Education colleges, City Technology Colleges, local authority alternative provision placements, and non-state-funded alternative provision providers, are not required to generate UPNs [15]. If a child’s first contact with education is through these providers, and a UPN is not generated for them, their CLA record cannot be linked to education datasets in the NPD. CLA and NHS datasets are not directly linked, instead children must first appear in the NPD to be linked to NHS records. Therefore, linkage across the CLA and NHS datasets depends on enrolment in state-funded schools.

Linkage between NPD and NHS datasets available in ECHILD, such as Hospital Episode Statistics, is carried out by NHS England. It is performed using deterministic algorithms that match the first four letters of forename, full surname, date of birth, postcode, and gender, producing a unique ECHILD ID. The resulting ECHILD linkage spine consolidates successful matches across education, health, and social care datasets, providing researchers with a longitudinal view of children’s interactions with these services [16]. Individuals can be linked across NHS, CLA and NPD data if they have a record in the Personal Demographics Service which includes anyone who has registered for NHS care and has an NHS number [17].

All identifiers are removed before researchers access the data. The National Data Opt-out allows individuals to prevent their health data from being used for research or planning beyond their own care; such records are excluded from research datasets, including ECHILD, in line with NHS Digital policy [18].

### 2.2. Population and follow-up period

We defined our population as Children Looked After with a UASC flag who have entered the CLA dataset for first time between April 1^sts^ 2005 and 31^st^ March 2021. Data were de-duplicated by keeping only the first episode of care, that is, the episode when they have first entered care, per child, appearing in the CLA for our follow-up period (April 1^st^, 2005 – 31^st^ March 2021).

### 2.3. Outcome variables and covariates

This study focused on two outcome variables: (1) any recorded enrolment in state-funded schools (2) any contact with NHS health services. 1) was measured by a link between CLA and NPD, and 2) by a link between CLA-NPD and PDS for a particular child.

All covariates (gender, age, ethnicity, region and placement type) were derived from the CLA dataset at first episode of care. Our covariates included gender, which is coded as a binary variable: male/female. The sex of children looked after should be reported as recorded on their birth certificate or gender recognition certificate [19].

Age at first episode of care was derived by calculating the difference between the reported date of birth and the start date of the first care episode and then recoded according to this cutoff. We recoded age into a binary variable (<16 years vs. ≥16 and <18 years). We set this age cut-off because evidence shows adolescents aged 16+ are less likely to attend state-funded schools for several reasons [20]. Education is no longer compulsory after 16 years of age. Furthermore, many move into Further Education (e.g., sixth form colleges), which are not classified as secondary schools. This distinction directly affects school enrolment and, in turn, linkage to the NPD.

Ethnicity was recoded using the DfE ethnicity codes from the NPD School Census. Social workers asked children to identify their own ethnicity and ethnic origin. They then recorded the pupil’s ethnicity by matching this description to the DfE’s Ethnic Origin Codes, which is a twenty-level categorical variable. These codes were collapsed into the main DfE categories, resulting in a five-level variable: “White,” “Black,” “Mixed,” “Asian,” and “Any Other.” This simplification captured the main ethnic groups and addressed low counts in many of the original sub-categories.

Region indicated Government Office Region of England (North East, North West, Yorkshire and the Humber, East Midlands, West Midlands, East of England, London, South East, and South West) of young person’s region of residence.

Finally, placement type referred to the type of placement provided by England’s children’s social care services. We recoded the detailed placement codes into broader categories, following the methodology outlined in McGrath-Lone [21]: Adoption, Foster (Kin), Foster (Stranger), Children’s Home, Residential Care Home, Residential School, Other Residential, With Parent, Independent, and Other. “Other residential” (also called semi-independent accommodation) and “Independent accommodation” are both forms of supported accommodation intended for older adolescents [22]. Our recoding differs from McGrath-Lone’s in some respects. Most notably, we included adoption within the “Other” category due to low numbers, and treated independent living (e.g., flats, lodgings, bedsits, B&Bs, or living with friends, with or without formal support) as a separate category, given its high prevalence among the UASC population.

More details on the way we have coded our socio-demographic and placement type variables can be found in Box 1 in the Appendix.

### 2.4. Statistical Analysis

We described the distribution of key characteristics within the UASC population in social care dataset, including age, gender, ethnic group, region and placement type. Then, we calculated overall linkage rates between the social care dataset and the school datasets, estimating how many UASC have enrolled in state-funded school and were therefore linked. We also calculated how many of those linked to the school datasets also appeared in the NHS datasets. Finally, we examined whether there were significant differences in linkage rates between the school and social care datasets and between the social care, school and NHS datasets depending on socio-demographic factors, region and placement type. We performed Chi-Square tests for differences in linkage across each covariate. We used a cut-off of *p* <0.01 to indicate statistical significance.

Statistical analyses were conducted in RStudio (version 4.3.3.) within the ONS Secure Research Service. In line with DfE confidentiality rules, counts are rounded to the nearest 10 and percentages are reported as whole numbers. The code used in this study is openly available at: https://github.com/RebeccaLangella/UASC_ECHILD_Cohort-_2005_2021-.git RL, KL and PH had access to the raw data during this study. The ECHILD database is made available for free for approved research based in the UK, via the ONS Secure Research Service. Enquiries to access the ECHILD database can be made by emailing. Researchers will need to be approved and submit a successful application to the ECHILD Data Access Committee and ONS Research Accreditation Panel to access the data, with strict statistical disclosure controls of all outputs of analyses□

## 3. Results

**Figure 1.**
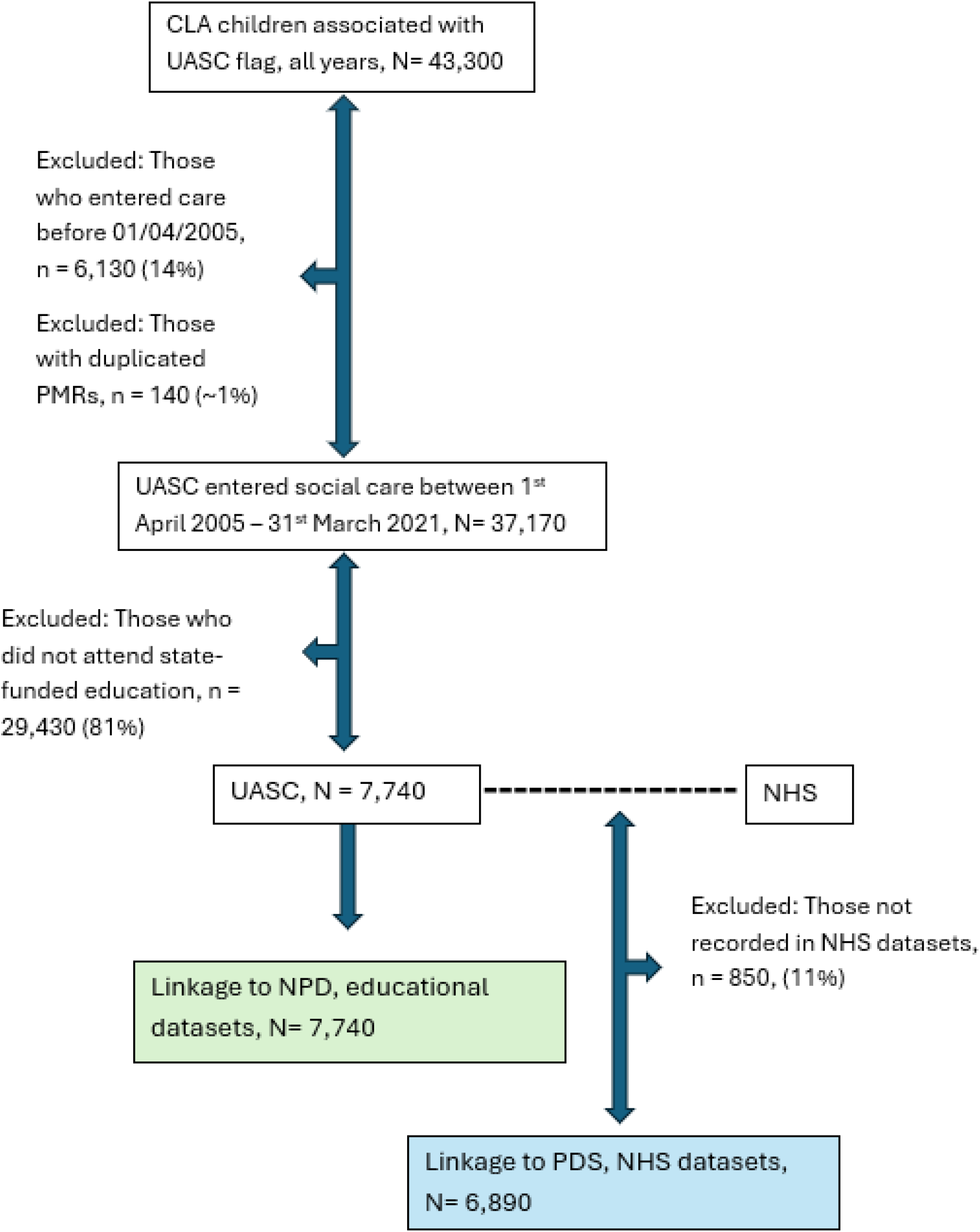
Flow Chart of cohort selection.

### 3.1. Socio-demographic, region of placement and placement of UASC in the CLA social care dataset

There were 37,170 UASC in the CLA dataset with a first record between 1^st^ April 2005 and 31^st^ March 2021, of which 32,570 (88%, Table 1) were male. Around 24,290 (66%) were aged 16–17-year-old (65%) at the start of their first episode of care. We also find that 10,310 (28%) were recorded as Black and 10,260 (27%) were recorded as Asian.

**Table 1.**
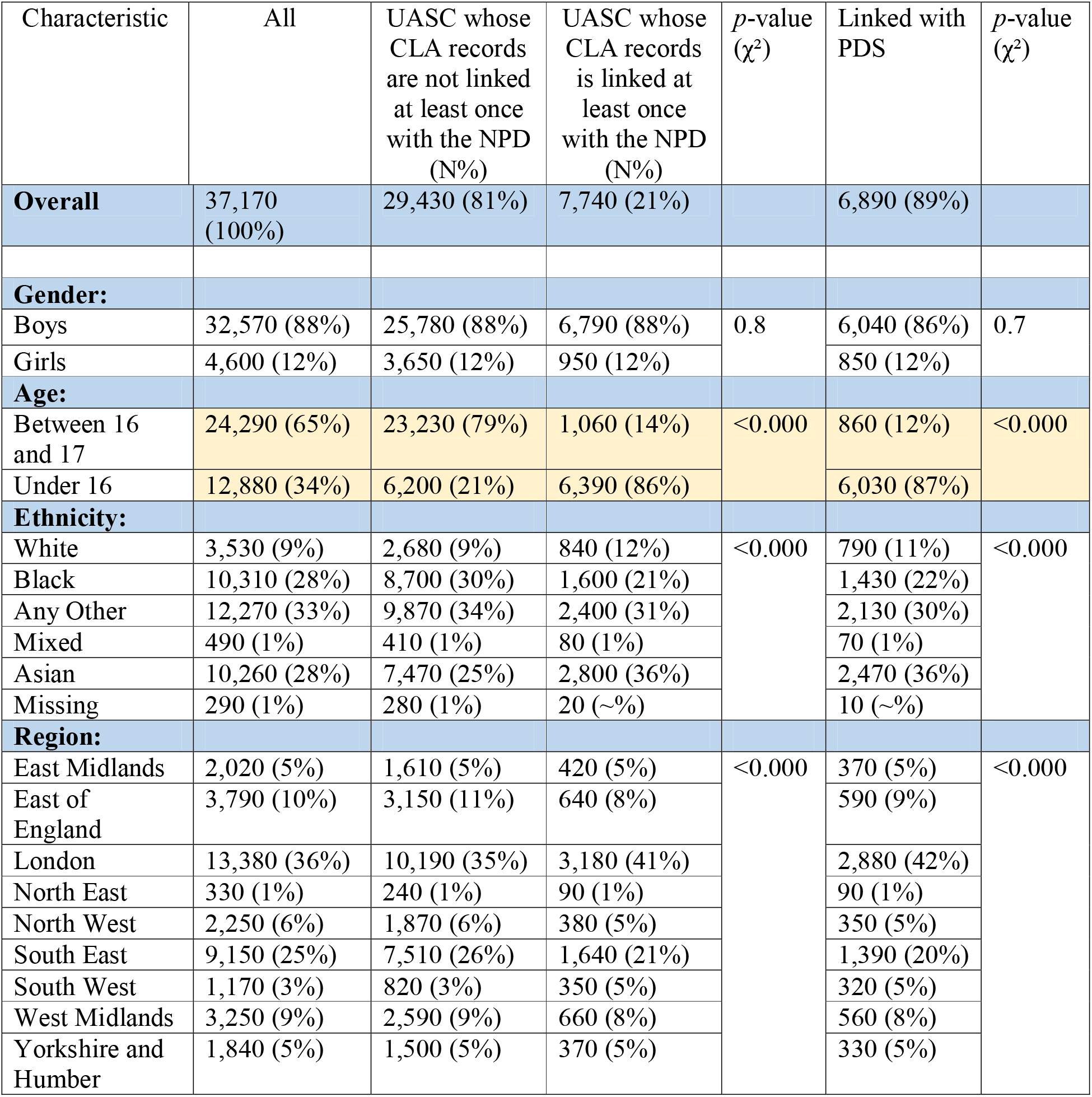
Sociodemographic characteristics and Pearson’s Chi-square test (χ^2^) UASC sample in the CLA linked and non-linked to the NPD and PDS, column percentages, 1^st^ April 2021 – 31^st^ March 2021.

| Characteristic | All | UASC whose CLA records are not linked at least once with the NPD (N%) | UASC whose CLA records is linked at least once with the NPD (N%) | p-value ($\chi^2$) | Linked with PDS | p-value ($\chi^2$) |
| --- | --- | --- | --- | --- | --- | --- |
| **Overall** | 37,170 (100%) | 29,430 (81%) | 7,740 (21%) |  | 6,890 (89%) |  |
| **Gender:** |  |  |  |  |  |  |
| Boys | 32,570 (88%) | 25,780 (88%) | 6,790 (88%) | 0.8 | 6,040 (86%) | 0.7 |
| Girls | 4,600 (12%) | 3,650 (12%) | 950 (12%) |  | 850 (12%) |  |
| **Age:** |  |  |  |  |  |  |
| Between 16 and 17 | 24,290 (65%) | 23,230 (79%) | 1,060 (14%) | <0.000 | 860 (12%) | <0.000 |
| Under 16 | 12,880 (34%) | 6,200 (21%) | 6,390 (86%) |  | 6,030 (87%) |  |
| **Ethnicity:** |  |  |  |  |  |  |
| White | 3,530 (9%) | 2,680 (9%) | 840 (12%) | <0.000 | 790 (11%) | <0.000 |
| Black | 10,310 (28%) | 8,700 (30%) | 1,600 (21%) |  | 1,430 (22%) |  |
| Any Other | 12,270 (33%) | 9,870 (34%) | 2,400 (31%) |  | 2,130 (30%) |  |
| Mixed | 490 (1%) | 410 (1%) | 80 (1%) |  | 70 (1%) |  |
| Asian | 10,260 (28%) | 7,470 (25%) | 2,800 (36%) |  | 2,470 (36%) |  |
| Missing | 290 (1%) | 280 (1%) | 20 (~%) |  | 10 (~%) |  |
| **Region:** |  |  |  |  |  |  |
| East Midlands | 2,020 (5%) | 1,610 (5%) | 420 (5%) | <0.000 | 370 (5%) | <0.000 |
| East of England | 3,790 (10%) | 3,150 (11%) | 640 (8%) |  | 590 (9%) |  |
| London | 13,380 (36%) | 10,190 (35%) | 3,180 (41%) |  | 2,880 (42%) |  |
| North East | 330 (1%) | 240 (1%) | 90 (1%) |  | 90 (1%) |  |
| North West | 2,250 (6%) | 1,870 (6%) | 380 (5%) |  | 350 (5%) |  |
| South East | 9,150 (25%) | 7,510 (26%) | 1,640 (21%) |  | 1,390 (20%) |  |
| South West | 1,170 (3%) | 820 (3%) | 350 (5%) |  | 320 (5%) |  |
| West Midlands | 3,250 (9%) | 2,590 (9%) | 660 (8%) |  | 560 (8%) |  |
| Yorkshire and Humber | 1,840 (5%) | 1,500 (5%) | 370 (5%) |  | 330 (5%) |  |

During their first episode of care, UASC were placed in three types of accommodation: 15,500 (42%, Table 2) were placed in stranger foster care, 8,810 (24%) were placed in independent placement, and 9,570 (26%) were placed in “Other Residential” placement. UASC were primarily resident in London and the Southeast; 13,390 (36%) and 9,150 (25%) respectively.

**Table 2.**
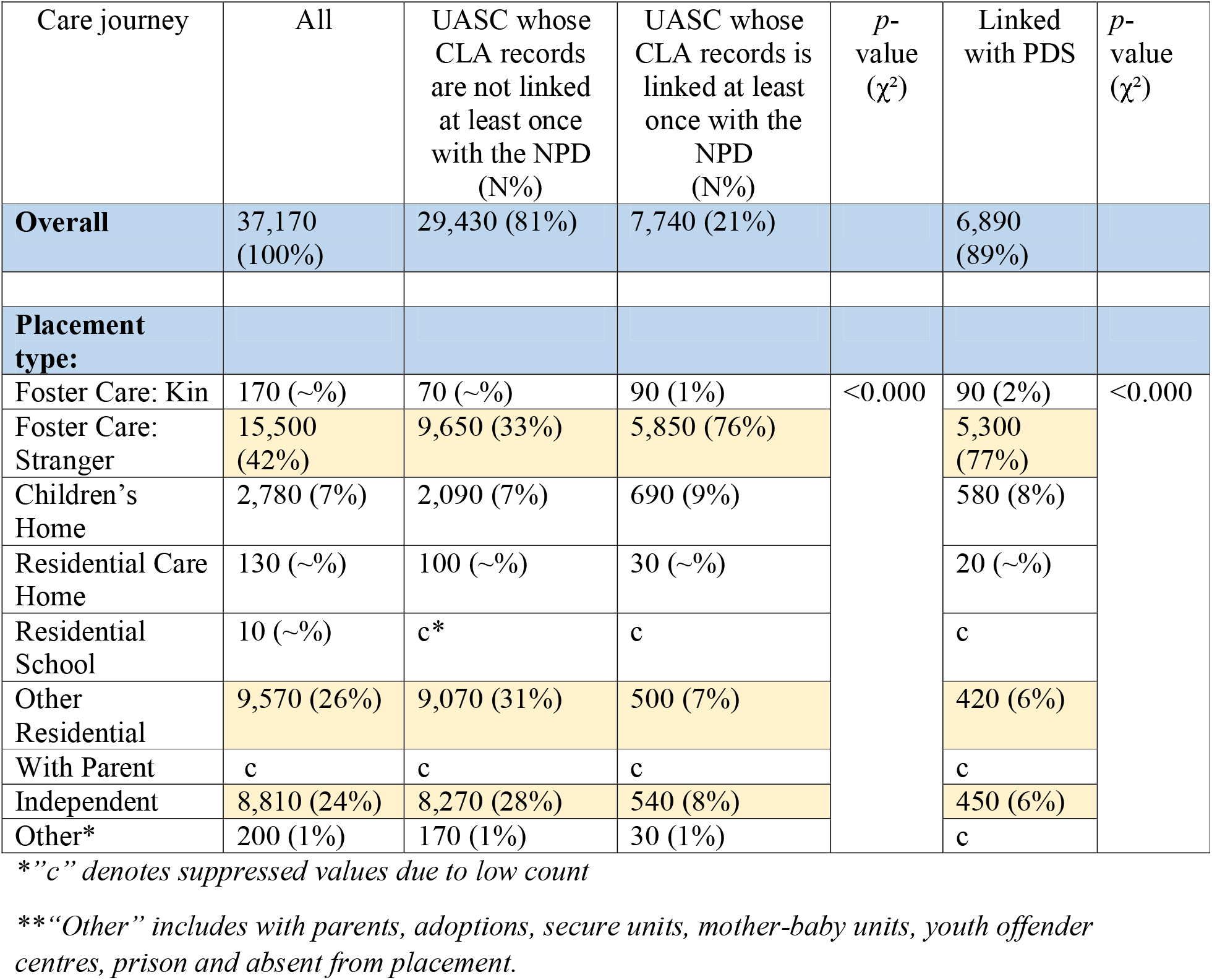
Placement type and Pearson’s Chi-square test (χ^2^) UASC sample in the CLA linked and non-linked to the NPD and PDS, column percentages, 1^st^ April 2021 – 31^st^ March 2021.

### 3.2. Differences in linkage rates: social care to educational datasets

Out of 37,170 UASC in the social care dataset, 7,740 were linked to the school datasets (21%; Figure 2). We found an association between linkage and age, ethnicity, region and placement type at first episode. Gender not found to be associated. Linkage to school datasets the differed significantly (*p* <0.001) according to age. Of the 24,290 UASC that are aged 16-17-years-old, only 1,060 (4%) were linked to the school datasets. In contrast, 5,770 (50%) of the 12,880 UASC under 16 years old were linked.

**Figure 2.**
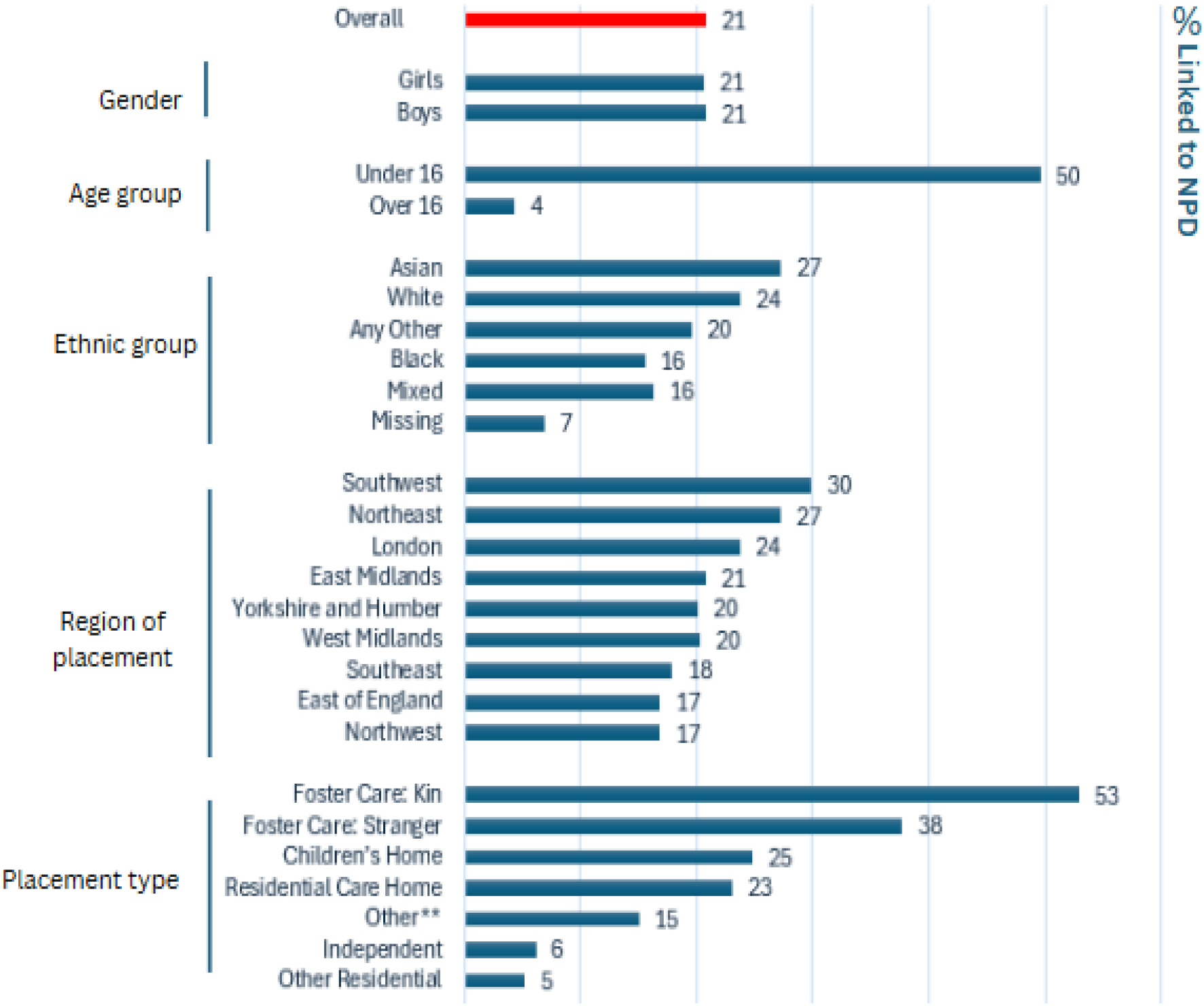
Percentage of UASC in the CLA linked to the NPD (1^st^ April 2005 – 31^st^ March 2021). *”Other” includes with parents, adoptions, secure units, mother-baby units, prison and absent from placement.

There were also significant differences (*p* <0.001) in linkage rates between ethnic groups: out of 10,310 Black UASC, only 1,600 (16%) were linked to the school datasets, whereas out of 10,260 Asian UASC, only 2,800 (27%) were linked. We also found that linkage rates varied significantly (*p* <0.000) by regions of England. Out of 1,170 UASC situated in the Southwest, 350 (30%) were linked. This is contrast to Northwest, were out of 2,250 (17%) having resided in the Northwest only 380 are linked.

Similarly, there were significant differences (*p* <0.001) in linkage between placement types, where only 500 (5%) of 9,570 UASC in Other Residential accommodation and 540 (6%) of 8,810 in independent accommodation were linked to the school datasets. This is in contrast with UASC in stranger foster care, where 5,650 (38%) of 15,500 UASC were linked.

### 3.3. Differences in linkage rates: school to NHS datasets

Linkage to the NHS datasets, mirrored a similar pattern to linkage with the school datasets. Of the 7,740 UASC linked to the school datasets, 6,890 were also linked to NHS datasets, with an 88% linkage rate between the social care-school and NHS datasets, but an overall linkage rate for UASC in the social care dataset to NHS datasets of 19% (Figure 3). We found an association between linkage and age, ethnicity, region and placement type at first episode. Gender not found to be associated. We found again that linkage rates (*p* <0.001) varied significantly by age. Out of the 11,260 UASC under 16 years, 6,030 (49%) were linked to the NHS datasets, whereas only 860 (4%) of 24,290 UASC aged 16-17-years old were linked to the NHS datasets.

**Figure 3.**
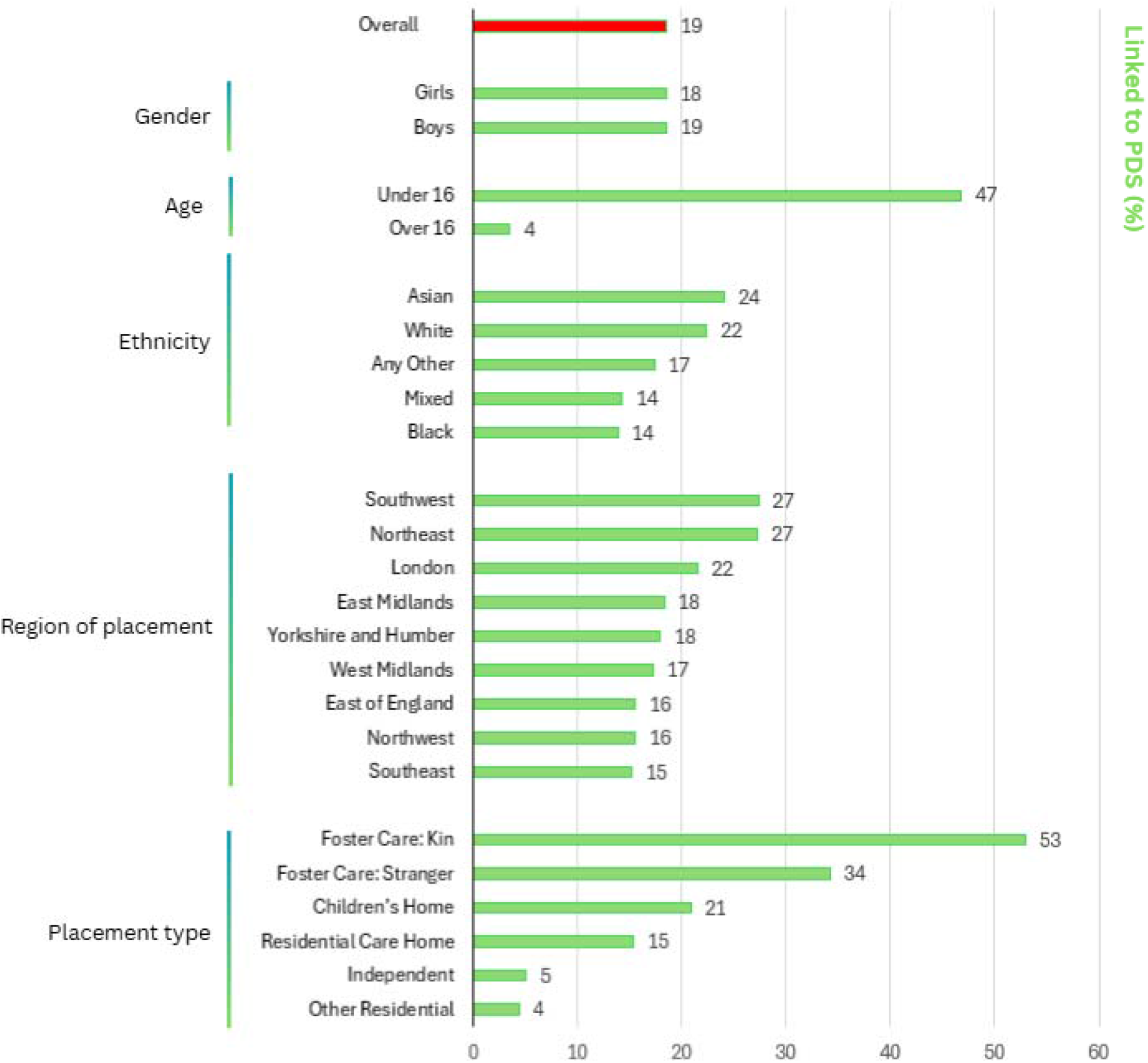
Percentage of UASC in the CLA linked to the NPD also linked to PDS (1^st^ April 2005 – 31^st^ March 2021). *” Other” category of placement type is not shown due to low count

We also found that linkage varied significantly (*p* <0.001) by ethnic groups: of 10,260 Asian UASC, 2,470 (24%) were linked, only 1,430 (14%) out of 10,310 Black UASC were linked. We found again that linkage rates varied significantly (*p* <0.001) by region of England, where out of 1,170 UASC situated in the Southwest, 320 (27%) were linked, with comparatively lower linkage rates in the South East, where only 1,390 (15%) out of 9,150 UASC were linked.

Finally, we found significant variation (*p* <0.001) in linkage rates between placement types. Only 420 (5%) of 9,570 UASC in Other Residential accommodation are linked to the NHS datasets. Similarly, only 450 (7%) of 8,810 in independent accommodation were linked, compared with 5,300 (35%) of 15,500 in foster care under the care of strangers.

## 4. Discussion

This data note provides the first national overview of linkage rates between social care, education, and healthcare datasets for UASC in England using the ECHILD dataset.

37,170 UASC were characterised in the social care dataset. The majority were male and aged 16-17 years. Most UASC were from Black or Asian ethnic backgrounds. Around 21% were linked to the educational dataset, and of those, 88% were also linked to the healthcare dataset, resulting in a 19% overall linkage rate from social care to healthcare. Linkage rates were notable higher for UASC aged under 16 years old compared with those aged 16-17 years old.

A major strength of this study is its national scope and the use of longitudinal administrative data spanning almost two decades. The creation of this cohort will enable unprecedented analysis of the intersections between social care, education, and healthcare trajectories for UASC, an area where UK-based quantitative evidence has been extremely limited. The ability to explore these domains in an integrated way offers new insights into systemic barriers and protective mechanisms for this highly vulnerable population.

However, several limitations must be acknowledged. Linkage to the NPD depends on enrolment in state-funded school. UASC are less likely than other children in care to be enrolled in state-funded schools, in part due to their older age (16-17 years old on average) and because they might also be receiving alternative forms of education that are not state-funded, including English language instruction, tutoring and bespoke programs [23]. Additionally, if a child’s first contact with education in England is through post-compulsory education, such as Further Education Colleges, they might not have a UPN and therefore cannot be linked to education datasets [24]. Further investigation is needed to determine whether their absence from educational datasets reflects non-attendance altogether, enrolment in alternative provision, or attendance at further education colleges. If they are not in education at all, these older adolescents might miss out on key opportunities for language learning and social integration [25].

Missed links among older UASC also limits the ability to evaluate the health implications of different care settings, particularly because placement assignment is closely tied to age.

UASC over 16 years old are overwhelmingly placed in supported accommodation designed for older children, as it is illegal to place children under 16 years in those placements [22]. These settings have been widely criticised by advocacy groups, including the Children’s Commissioner, for failing to provide adequate safeguarding, access to education, healthcare, and legal support (24, 25). It is essential that data collection efforts do not inadvertently reinforce these inequalities, and that datasets are inclusive enough to capture the experiences of older UASC who are not enrolled in state-funded school.

A substantial proportion of UASC (32%) are categorised as “Any other ethnicity,” likely including children from Middle Eastern, North African, and other backgrounds not explicitly defined in the standardized ethnic categories provided by the DfE. Ethnicity is self-reported or reported by a carer and recorded using standardised codes [26], providing only a general indication of ethnic background. Nevertheless, our analysis identified that UASC from Black ethnic groups were underrepresented in the linked cohort compared to the full UASC cohort in the social care data, highlighting the need for further investigation.

Finally, all descriptive variables, including region and placement type, refer only to the first recorded entry in the social care dataset. As a result, changes in placement or care experiences over time were not captured in this paper, although it will be possible to do so in future research using this cohort. Despite these limitations, the socio-demographic characteristics, regional distribution, and placement types observed in this study closely mirror those reported in the Department for Education’s Children Looked After statistics. For instance, the 2020 annual CLA return indicates that 90% of UASC were male, 13% were under 16 years old, 50% were in supported accommodation (independent or semi-independent), and 48% were in foster care, with similar distributions observed from 2021 to 2024 [27]. These patterns align with our findings and suggest consistency over time. However, prior to 2020, DfE statistics did not provide disaggregated data by year of entry, including the number of UASC starting care annually, highlighting the need for retrospective, dedicated research on this population.

## 5. Conclusion

We estimated that fewer than one quarter of UASC are enrolled state-funded school, and thus to be linkable to health data within the ECHILD dataset. The reliance on school enrolment as a prerequisite for linkage to both educational and health data significantly limits the representation of UASC in the dataset for research into health and education outcomes. Despite limitations, we have identified a sample of 7,740 UASC linked to education data across our study period (2005 – 2021) and 6,890 that are linked to both educational and health data. This population provides a foundation for longitudinal analysis of educational and health outcomes of UASC, offering a unique opportunity to explore their relationship with socio-demographic factors and care experiences. It represents an unprecedented resource and a major advancement in understanding the needs and long-term outcomes of UASC nationwide.

## Data Availability

The ECHILD database is made available for free for approved research based in the UK, via the ONS Secure Research Service. Enquiries to access the ECHILD database can be made by emailing. Researchers will need to be approved and submit a successful application to the ECHILD Data Access Committee and ONS Research Accreditation Panel to access the data, with strict statistical disclosure controls of all outputs of analyses

## Ethics and consent□

Permissions to use linked, de-identified data from Hospital Episode Statistics and the National Pupil Database were granted by DfE (DR200604.02B) and NHS Digital (DARS-NIC-381972). Ethical approval for the ECHILD project was granted by the National Research Ethics Service (17/LO/1494), NHS Health Research Authority Research Ethics Committee (21/SW/0159) and UCL Great Ormond Street Institute of Child Health’s Joint Research and Development Office (20PE06). Patient consent was not required to use the deidentified data in this study.□

## Appendix

**Box 1.**
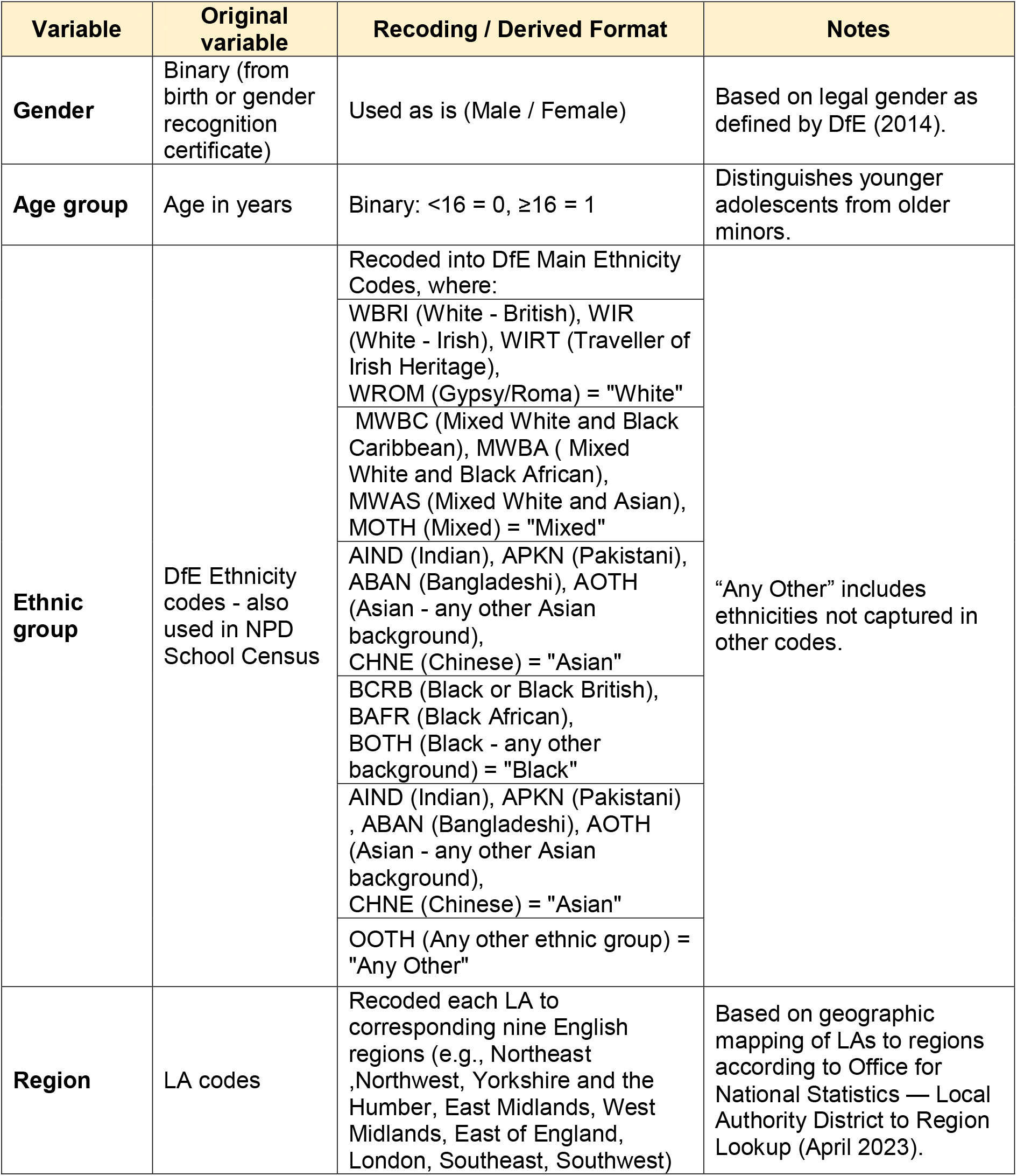
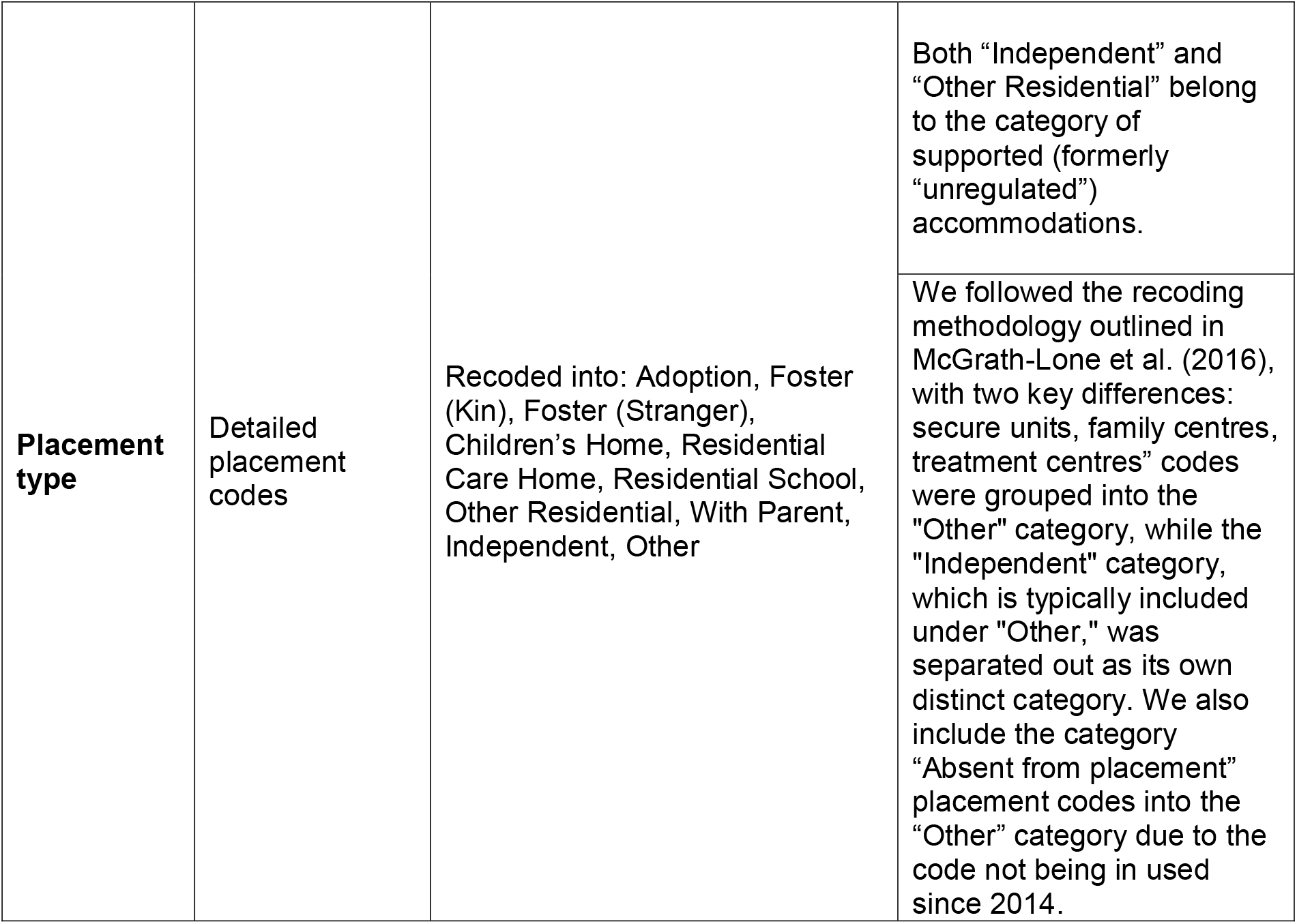
Recoding of variables used in analysis for Objective 1A.

## Bibliography

1. UK Home Office. Immigration Rules: Appendix asylum – Unaccompanied asylum-seeking children (Paragraph 352ZD). In: Immigration Rules (archived version, 1 October 2019). GOV.UK; 2019. Available from: https://assets.publishing.service.gov.uk/media/5f68f04f8fa8f5076adc46c1/Immigration_Rules_-_Archive_01-10-2019.pdf

2. Dorling R, MacLachlan M, Trevena L. Analysis of care and education pathways of refugee and asylum-seeking children in care. Oxford: Oxford University Research Archive; 2017. Available from: https://ora.ox.ac.uk/objects/uuid%3Af15a518e-141e-4b9f-b779-7b1de94d195d/files/md884abbaaee0164e5fd6c46f4705bc09

3. Department for Education. Children looked after in England including adoptions: 2024. GOV.UK; 2024. Available from: https://explore-education-statistics.service.gov.uk/find-statistics/children-looked-after-in-england-including-adoptions/2024

4. Department for Education. Children looked after in England including adoptions: 2019. GOV.UK; 2019. Available from: https://assets.publishing.service.gov.uk/media/5de7a998e5274a06d662b19f/Children_looked_after_in_England_2019_Text.pdf

5. Yim SH, Said G, King D. Practical recommendations for addressing the psychological needs of unaccompanied asylum-seeking children in England: a literature and service review. Clin Child Psychol Psychiatry. 2025;30(2):245–63. doi:10.1177/13591045241252858. Available from: https://journals.sagepub.com/doi/10.1177/13591045241252858

6. Home Office. Understanding asylum-seeker and asylum-route refugee vulnerabilities, needs and support. UK Government; 2022. Available from: https://www.gov.uk/government/publications/understanding-asylum-seeker-and-refugee-vulnerabilities-and-needs-2022/understanding-asylum-seeker-and-asylum-route-refugee-vulnerabilities-needs-and-support-2022

7. Mulcaire J, Smetham D, Holt L, et al. Impact of the asylum determination process on mental health in the UK and EU+: a systematic review and thematic synthesis. BMJ Public Health. 2024;2:e000814. doi:10.1136/bmjph-2023-000814. Available from: https://bmjpublichealth.bmj.com/content/2/2/e000814

8. Shahzad A, Katona C, Glover N. The psychological impact of spending a prolonged time awaiting asylum. Eur J Psychotraumatol. 2025;16(1):2506189. doi:10.1080/20008066.2025.2506189. Available from: https://www.tandfonline.com/doi/full/10.1080/20008066.2025.2506189

9. UK. Education for refugee and asylum seeking children: Access and equality in England, Scotland and Wales. Refugee Support Network; 2018. Available from: https://www.unicef.org.uk/wp-content/uploads/2018/09/Access-to-Education-report-PDF.pdf

10. O’Higgins A, Ott EM, Shea MW. What is the impact of placement type on educational and health outcomes of unaccompanied refugee minors? A systematic review of the evidence. Clin Child Fam Psychol Rev. 2018;21:354–65. doi:10.1007/s10567-018-0256-7. Available from: https://link.springer.com/article/10.1007/s10567-018-0256-7

11. Hutchinson J, Reader M. The educational outcomes of refugee and asylum-seeking children in England: an experimental methodology for analysing attainment, absence and exclusions. Education Policy Institute; 2021. Available from: https://dera.ioe.ac.uk/id/eprint/39826/

12. Sebba J, Berridge D, Luke N, Fletcher J, Bell K, Strand S, et al. The educational progress of looked after children in England: linking care and educational data. London: Nuffield Foundation; 2015. Available from: http://www.nuffieldfoundation.org/educational-progress-looked-afterchildren

13. Ramzan F, McGrath-Lone L, Gilbert R, Blackburn R, Ruiz Nishiki M, Lilliman M, et al. Education & Child Health Insights from Linked Data (ECHILD): an introductory guide for researchers (Version 2.1.4) [User guide]. London: UCL Great Ormond Street Institute of Child Health; 2023. Available from: https://docs.echild.ac.uk/

14. Jay MA, McGrath-Lone L, Gilbert R. Data resource: The National Pupil Database (NPD). Int J Popul Data Sci. 2019;4(1):1101. doi:10.23889/ijpds.v4i1.1101. Available from: https://doi.org/10.23889/ijpds.v4i1.1101

15. Department for Education. Unique pupil numbers (UPNs): a guide for schools and local authorities (Version 1.2). GOV.UK; 2019. Available from: https://assets.publishing.service.gov.uk/media/5cfa739a40f0b663fd865a6a/UPN_Guide_1.2.pdf

16. Libuy N, Harron K, Gilbert R, Caulton R, Cameron E, Blackburn R. Linking education and hospital data in England: linkage process and quality. Int J Popul Data Sci. 2021;6(1):1671. doi:10.23889/ijpds.v6i1.1671. Available from: https://doi.org/10.23889/ijpds.v6i1.1671

17. NHS. What is an NHS number? NHS; 2023 May 16 [cited 2025 Oct 17]. Available from: https://www.nhs.uk/using-the-nhs/about-the-nhs/what-is-an-nhs-number/

18. NHS Digital. National Data Opt-Out: Operational Policy Guidance Document. NHS Digital; 2021. Available from: https://digital.nhs.uk/services/national-data-opt-out/operational-policy-guidance-document

19. Department for Education. Children looked-after by local authorities in England: guide to the SSDA903 collection 1 April 2024 to 31 March 2025 (Version 1.3). GOV.UK; 2025. Available from: https://assets.publishing.service.gov.uk/media/67ee564ae9c76fa33048c702/Children_looked-after_by_LAs_in_England_2024-25_Guide_Version_1_3.pdf

20. Department for Education. Participation headline data from “Participation in education, training and employment age 16 to 18”. Explore Education Statistics; 2022. Available from: https://explore-education-statistics.service.gov.uk/data-tables/permalink/6df4d3ea-1a0f-4d67-9932-5f346f7de40b

21. McGrath-Lone L, Harron K, Dearden L, Nasim B, Gilbert R. Data resource profile: Children Looked After Return (CLA). Int J Epidemiol. 2016;45(3):716–717f. doi:10.1093/ije/dyw117. Available from: https://doi.org/10.1093/ije/dyw117

22. Department for Education. Guide to the Supported Accommodation Regulations including Quality Standards. GOV.UK; 2023. Available from: https://assets.publishing.service.gov.uk/media/6514400088281e000db4e965/Guide_to_the_supported_accommodation_regulations_including_quality_standards.pdf

23. O’Higgins A. Analysis of care and education pathways of refugee and asylum-seeking children in care in England: implications for social work. Int J Soc Welf. 2019;28:53– doi:10.1111/ijsw.12324. Available from: https://onlinelibrary.wiley.com/doi/10.1111/ijsw.12324

24. Children’s Commissioner for England. Unregulated: children in care living in semi-independent accommodation. London: Children’s Commissioner; 2020. Available from: https://www.childrenscommissioner.gov.uk/resource/unregulated/

25. Children’s Commissioner. Looked After Children not in school: data report. London: Children’s Commissioner; 2023 May. Available from: https://assets.childrenscommissioner.gov.uk/wpuploads/2023/05/cc-lac-not-in-school.pdf

26. Department for Education. Children looked after by LAs in England 2024–25: guide (Version 1.3). GOV.UK; 2025. Available from: https://assets.publishing.service.gov.uk/media/67ee564ae9c76fa33048c702/Children_looked-after_by_LAs_in_England_2024-25_Guide_Version_1_3.pdf

27. Department for Education. Children looked after in England including adoptions: reporting year 2024. GOV.UK; 2024 Nov 14. Available from: https://explore-education-statistics.service.gov.uk/find-statistics/children-looked-after-in-england-including-adoptions/2024

